# Profile of the pregnant woman with Gestational Diabetes Mellitus in a private hospital in Joinville, Brazil

**DOI:** 10.1101/2022.12.10.22283277

**Authors:** João Pedro Ribeiro Baptista, Matheus Augusto Schulz, Lehana Fleith, Israel Kitzberger, Wesley Feitosa Hilário, Dieter Alisson Neumann, Júlia Opolski Nunes da Silva, Rodrigo Ribeiro e Silva, Iramar Baptistella do Nascimento, Jean Carl Silva

## Abstract

**Objective:** To analyze the maternal-fetal characteristics of patients with gestational diabetes mellitus (GDM) attended in a private service.

**Materials and Methods:** This is a cross-sectional study. Data collection was carried out from the medical records of patients diagnosed with GDM at Centro Hospitalar UNIMED, in Joinville city, between 2011 and 2017. Maternal-fetal characteristics, therapeutics and complications of pregnant women diagnosed with GDM were analyzed.

**Results:** 515 patients with GDM were reported. Of the maternal characteristics, the mean age was (31.77), BMI (27.82). The proportion of obese pregnant women was (55.6%), normal (42.4%) and low weight (2%). The preference for cesarean section was (81.4%) and Gestational Age (GA) of GDM diagnosis (26.85), with higher diagnosis in the third trimester (65.2%) than second (29.3%) and first (5.3%). Incidences of hypertension (7.4%), preeclampsia (0.6%) and pregnant women with previous abortions (16.5%) were found. The mean glycemic profile showed HbA1C (5.26), estimated glucose (104.30) and FPG fasting (86.38), in the first hour (144.46) and in the second hour (64.15). Treatment with diet and exercise (38.3%), metformin (40%), insulin (14.5%) and combined (7.8%). Regarding newborns, the mean weight was (3.123.46) and the GA at birth (37.46). Regarding the percentile, AGP (71.8%), SGP (28.2%) and no cases of LGP. In terms of adverse outcomes, hyperbilirubinemia was evidenced in (25.6%), prematurity (10.9%) and fetal hypoglycemia (9.9%).

**Conclusion:** Pregnant women diagnosed with GDM had a higher GA at diagnosis in the third trimester and more cesarean sections. Among the adverse outcomes related to newborns, there were more cases of hyperbilirubinemia, prematurity and fetal hypoglycemia.

## Introduction

Gestational Diabetes Mellitus (GDM) can be defined as any level of glucose intolerance, resulting in a state of variable hyperglycemia, whose onset occurred and was diagnosed during pregnancy. The pathophysiology is due to the elevation of counter-regulatory hormones, mainly insulin, due to risk factors, either genetic or environmental, and to the stress generated by pregnancy. The incidence varies according to the studied population and according to the criteria used, being estimated between 3% and 7% in Brazil, with about 150,000 new cases per year (1).

GDM increases the risk of numerous perinatal outcomes, including maternal and perinatal mortality, premature rupture of membranes, preterm birth, preeclampsia, macrosomia, abortion, ICU admissions, jaundice, congenital malformations and neonatal hypoglycemia (2,3). It is worth noting that 15% to 50% of women with GDM have a chance of developing diabetes or glucose intolerance post-gestation (4). Therefore, reducing these cases would impact the incidence of clinical diabetes.

In the long term, intrauterine hyperglycemia increases the risk of metabolic syndrome, such as type 2 diabetes, dyslipidemia and obesity, hypertension, cardiovascular diseases and some types of cancer in adulthood (GG). Such consequences reinforce the importance of a multidisciplinary team as part of prenatal care, centered on the individual needs of each woman, clarifying doubts about the diagnosis, treatment and repercussions of the disease on herself and on her concept, increasing adherence to therapeutic indications and, consequently, improving results (5).

In the context of GDM treatment, the primary approach is diet and physical exercise, being the first measures to be taken, and, in case of failure, association of oral hypoglycemic agents, insulin or combinations of these. Such treatment should be adapted to the characteristics and individual needs of each patient and has proved to be effective in reducing maternal glycemic levels and, consequently, improving fetal evolution.

It is worth noting the importance of new studies on GDM for a better understanding of development, frequency, risk factors, treatment and prevention of this disease, contributing to the formulation of better practices. Thus, this study aims to analyze the maternal-fetal characteristics related to GDM and its possible adverse outcomes.

## Methodology

This is a cross-sectional study. The population comprised pregnant women diagnosed with GDM, who were attended from January 2011 to December 2017 in the High Risk Service of the Centro Hospitalar Unimed (CHU), located in Joinville (SC), Brazil. All had their delivery at the same service.

Inclusion criteria were pregnant women over 18 years old, with single gestation, who gave birth at the Centro Hospitalar UNIMED between January 2011 and December 2017, and patients diagnosed with GDM. Pregnant women with incomplete data or fetuses who evolved to fetal death were excluded.

Maternal-fetal characteristics evaluated in the study were age, BMI, gestational age at GDM diagnosis, presence of hypertension, glycosylated hemoglobin, previous cesarean sections and therapeutic options (diet, metformin or insulin). Fetal characteristics evaluated were capurro, weight, normal weight, HGT 1 and 2, APGAR of 1st and 5th minute, size classification (SGA, AGA and LGA), besides adverse outcomes (prematurity, neonatal ICU admission, respiratory distress, hyperbilirubinemia, hypoglycemia, low APGAR of 1st and 5th minute).

Information regarding patient characteristics, treatments involved and newborn outcomes were collected by the medical staff. The obesity classification used, through the Body Mass Index (BMI), follows the standards established by the World Health Organization (WHO), which are classified as low weight (<18.5), normal (18.5-24.9), overweight (25.0-29.9) and obesity (≥ 30.0).

Pregnant women included in this group were diagnosed according to the guidelines of the Brazilian Diabetes Society (SBD) which are the same as those of the World Health Organization (WHO). Screening for gestational ages (GA) below 20 weeks was performed through the fasting glucose test. GDM diagnosis was made when the result was between 92 and 125 mg / dl. From 24 weeks of gestation, all underwent the oral glucose tolerance test (OGTT). The reference values for GDM are: fasting glycemia ≥ 92, glycemia after 1h ≥ 180 mg / dl or after 2h ≥ 153 mg / dl, being that any of the points being altered in the curve already makes the diagnosis. (6)

The routine used during each medical consultation for the definition of the therapeutic proposal is based on a clinical-laboratory score. It is composed of five parameters with scores ranging from -2 to +2. The criteria evaluated are: fasting glycemia, postprandial glycemia, fetal abdominal circumference, maternal BMI and GA in the consultation. Thus, four recommendations are made according to the total score of the factors added. Scores below zero indicate a new consultation with a nutritionist, between zero and two, maintenance of the diet and physical exercise, between two and four, introduction of oral hypoglycemic and when greater than four indicates administration of insulin plus oral drug. (7)

The project was approved under the number CAAE 77220417.7.0000.5366 by the Research Ethics Committee (CEP) of the University of the Region of Joinville - UNIVILLE, Joinville (SC), Brazil. The study only began after the CEP approval opinion and followed in its development the requirements of Resolution 466/12 of the National Council of Health of the Ministry of Health.

All the information obtained was entered into the Microsoft Excel version 2016 software and later analyzed through the Statistical Package for the Social Science (SPSS) version 21.0. Quantitative variables were presented by means and standard deviations and qualitative variables by absolute and relative frequencies.

## Results

515 postpartum women were diagnosed with GDM. Regarding maternal characteristics (Table 1), women diagnosed on average were 31 years old, their average height and weight were 1.62m and 73.8kg, thus obtaining 10 postpartum women with underweight BMI, 216 with normal weight and 283 obese. 38 pregnant women had hypertension and 3 developed gestational diabetes hypoglycemia. Previous abortion was present in 85 women and the average gestational age of GDM detection was 26.8 weeks. Regarding the trimester of GDM diagnosis, 27 pregnant women were diagnosed in the first trimester, 149 in the second trimester and 332 in the third trimester.

**Table 1:** Maternal characteristics of patients diagnosed with GDM:

|  | Postpartum women diagnosed with GDM<br>(N=515) |
| --- | --- |
| Maternal age | 31,77 (4,9) |
| Height | 1,62 (0,0) |
| Weight | 73,84 (19,5) |
| BMI | 27,82 (6,164) |
| Low weight | 10 (2,0) |
| Normal | 216 (42,4) |
| Obese | 283 (55,6) |
| Arterial Hypertension | 38 (7,4) |
| Hypertensive Disease of Pregnancy | 3 (0,6) |
| Previous abortion | 85 (16,5) |
| GA at Diagnosis | 26,85 (7,6) |
| Trimester of diagnosis |  |
| 1st trimester | 27 (5,3) |
| 2nd trimester | 149 (29,3) |
| 3rd trimester | 332 (65,2) |

Mean and standard deviation, absolute numbers and percentages; BMI – Body Mass Index; GA – Gestational Age;

Regarding the glycemic profile of the patients with GDM (Table 2), the average value of glycosylated hemoglobin was 5.26, within the normal range. In addition, the estimated average glycemia was 104.3. As for the FPG, the average value was 86.3, the first hour curve was 144.4 and the second hour was 112.7. The value of glucose HGT 1 for the patients was on average 64.1 and glucose HGT 2 was 63.3. Thus, 197 patients treated GDM with diet and exercise, 206 received metformin for treatment, 74 used insulin and 40 patients made the association of metformin and insulin to treat GDM.

**Table 2:** Glycemic Profile and Treatment of Patients Diagnosed with GDM

|  |  |
| --- | --- |
|  | Postpartum women diagnosed with GDM<br>(N=515) |
| Glycemic profile |  |
| Hb A1c | 5,26 (0,4) |
| Estimated glycemia | 104,30 (11,6) |
| OGTT |  |
| Fasting | 86,38 (9,7) |
| 1st hour | 144,46 (36,1) |
| 2nd hour | 112,77 (31,1) |
| Glucose HGT 1 | 64,15 (21,4) |
| Glucose HGT 1 | 63,38 (22,8) |
| Treatment: |  |
| Diet and exercise | 197 (38,3) |
| Metformin | 206 (40,0) |
| Insulin | 74 (14,5) |
| Metformin and Insulin | 40 (7,8) |

Mean and standard deviation, absolute numbers and percentages; HbA1c - Glycosylated Hemoglobin; TOTG - Oral Glucose Tolerance Test.

In this regard, regarding the characteristics of the newborn of mothers with GDM (Table 3), the average weight of the newborn was 3,123.4g and the gestational age of birth was 37.4 weeks. The Apgar of the first minute was 7.9 and the fifth minute was 8.8. As for the abdominal circumference, the value was 327.3mm and the head circumference was 332.5mm. About the delivery route, 95 pregnant women had normal delivery and 419 had cesarean delivery. In this context, the fetal percentile average was 83.2, thus there were no SGA babies, the number of AGA babies was 370 and 145 newborns were LGA.

**Table 3:**
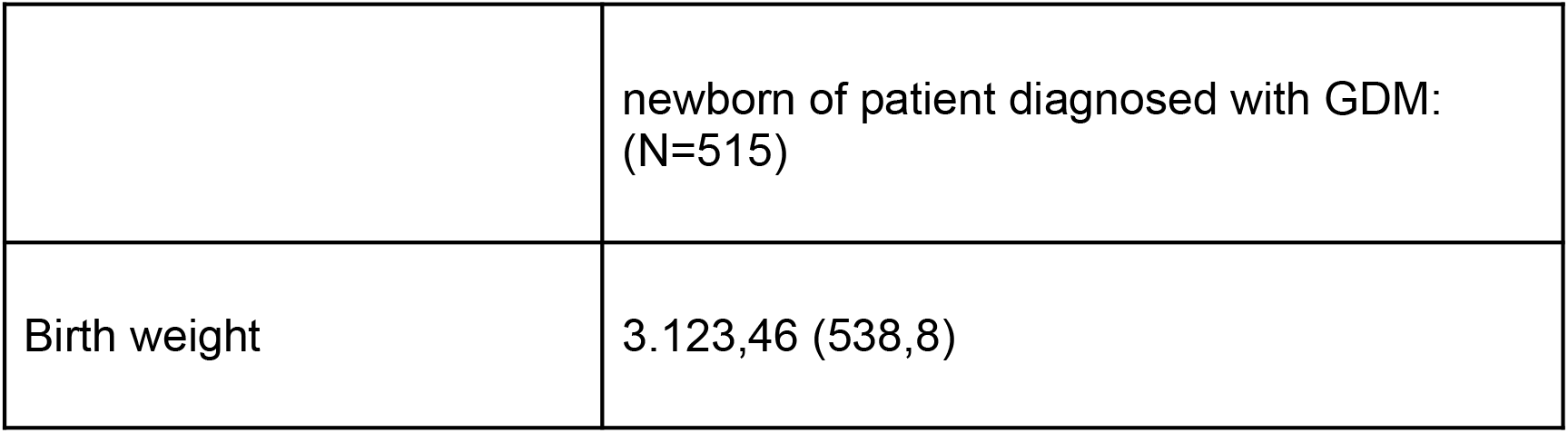
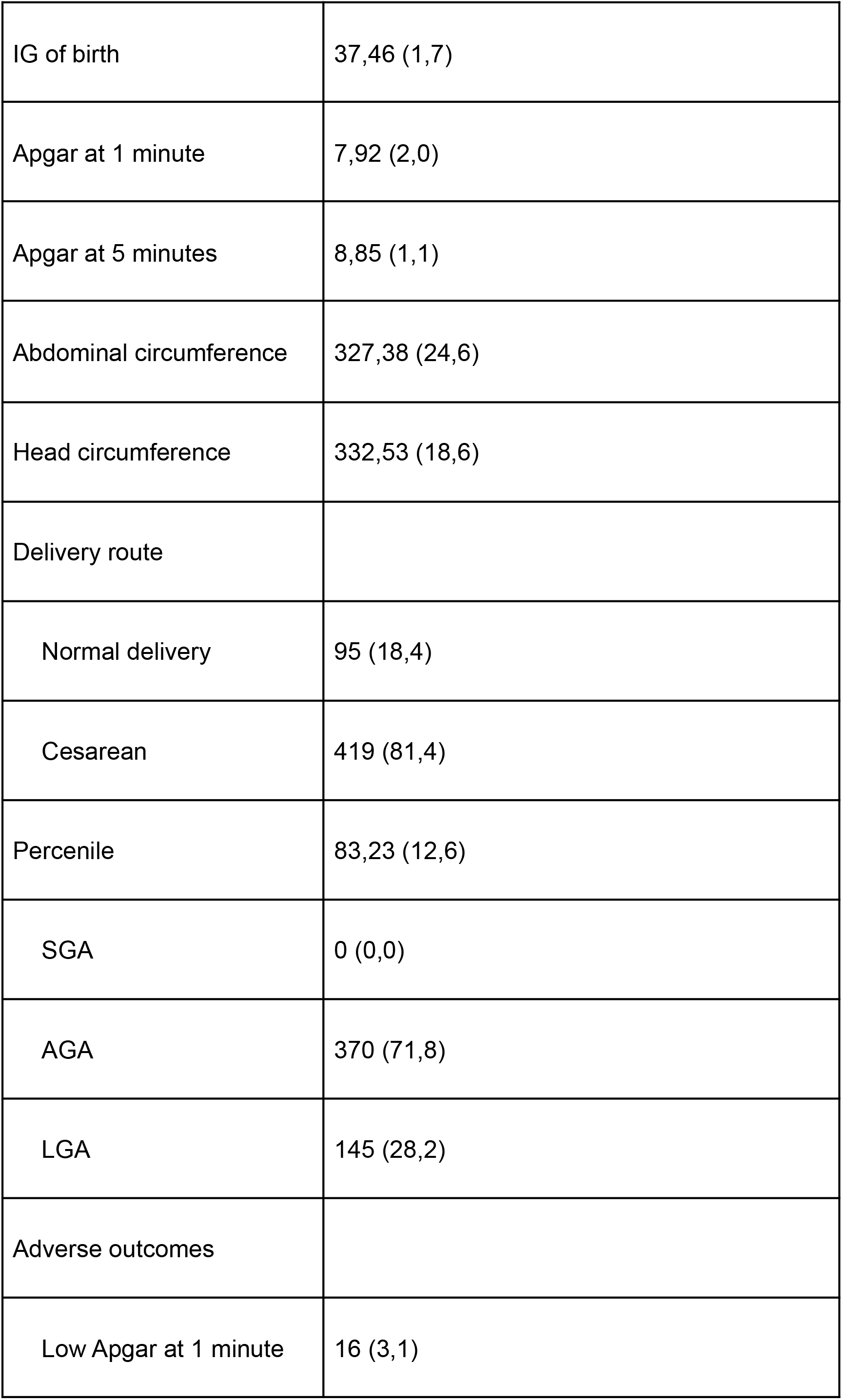
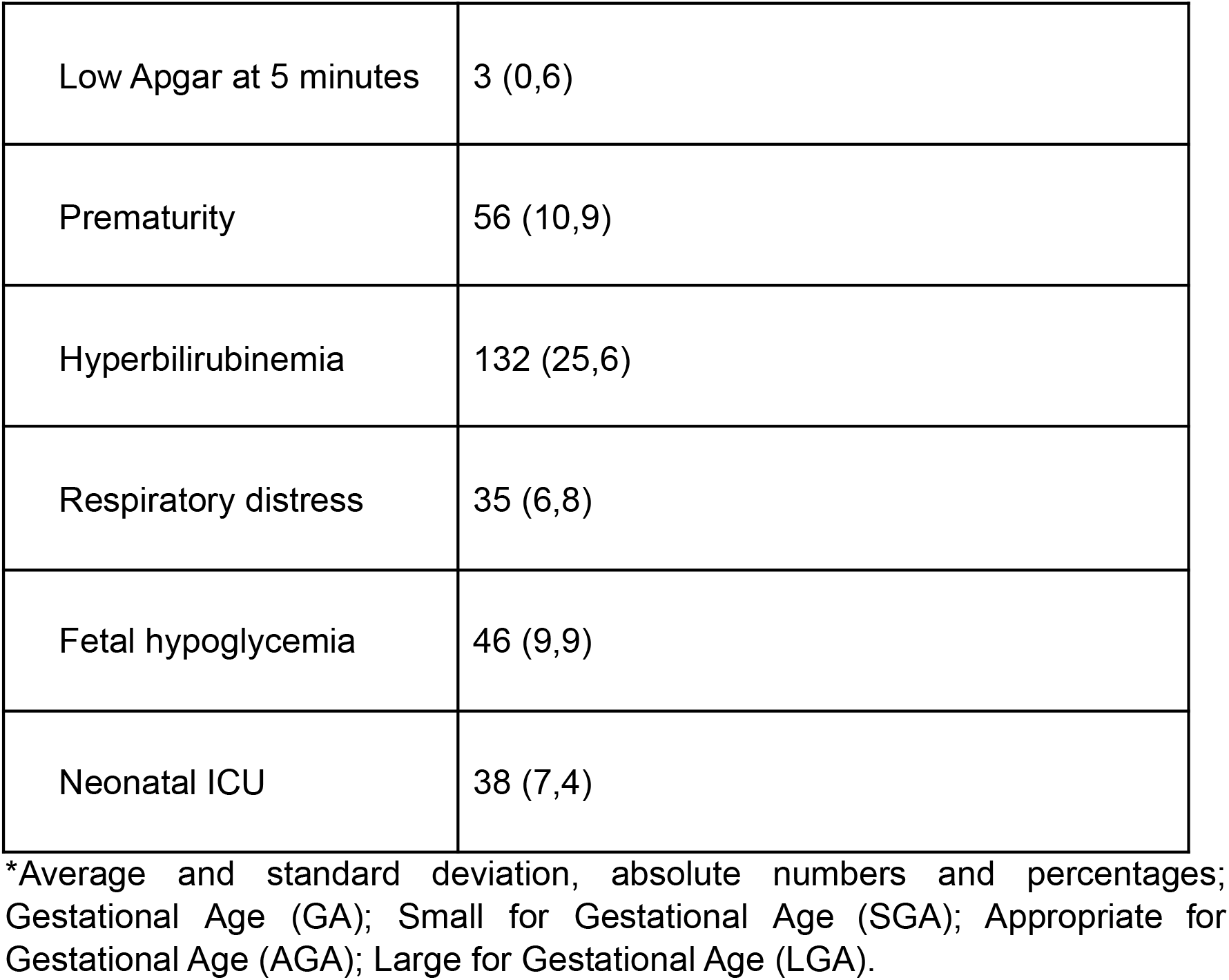
Characteristics of the newborn of patient diagnosed with GDM:

Regarding the adverse outcomes, 16 (3.1%) had a low apgar at the first minute and 3 had a low apgar at the fifth minute. In addition, 56 (10.9%) newborns were premature and 132 (25.6%) had hyperbilirubinemia. Babies who presented respiratory distress were 35 (6.8%), 46 (9.9%) suffered fetal hypoglycemia and 38 (7.4%) newborns needed neonatal ICU.

## Discussion

Given the importance of the diagnosis and control of GDM and its consequences, both maternal and fetal, the present study aimed to analyze the maternal-fetal characteristics related to GDM and its possible adverse outcomes.

A study conducted in the city of Fortaleza including all patients with GDM diagnosed hospitalized in the period of 2006 in a high-risk obstetric sector brought similar results regarding age: 60.7% of patients with GDM presented an age superior to 30 years, corroborating with the finding of the present study that indicated an average of 31 years of age for women with the same diagnosis (8). Another study carried out at the University Hospital of the Federal University of Maranhão pointed out an average age of 32.7 years (9). In this sense, the relationship between advanced age and risk of GDM (3) is confirmed. Some studies defined progressive risk for GDM from the age of 25 years (10). In Australia, a risk for GDM was found four times higher in women aged 35 to 39 and six times in those aged over 40 (11).

The maternal obesity present in 56% of the studied population confirms the findings of a study conducted in a group of patients at Hospital das Clínicas da UFMG with GDM diagnosis, which found only 30% of patients with pre-conceptual adequate BMI (5). Another study conducted in Recife found 189 (30.6%) pregnant women with GDM diagnosis with excess weight and 154 (25.0%) with obesity (12), thus, more than half of the pregnant women with GDM diagnosis presented excess weight or obesity. Such national studies reinforce the relationship of predisposition to GDM in obese pregnant women, already known in the literature (13,14).

Considering the high prevalence of obesity among pregnant women with GDM diagnosis, the concomitance of the diagnosis of hypertension or occurrence of DHEG in these patients is understood. This is because pregnant women who present excessive weight gain during pregnancy or who already start this period with overweight or obesity tend to develop, in addition to GDM, especially at the end of pregnancy, the Specific Hypertension Disease of Pregnancy (15). In addition, well-established studies of Cardiology confirm that obesity, defined as Body Mass Index (BMI)> 30 kg/ m2, is an important risk factor for arterial hypertension (16,17).

With regard to the obstetric history, it is noteworthy that 16.5% reported a previous abortion, a data very close to the prevalence of the report of spontaneous abortion in the general female population, which is 14% according to a Brazilian study on abortion with a demographic focus (18). Conversely, a study on the profile of mothers and newborns in the presence of GDM resulted in 35.2% of previous abortion (8), highlighting that the bibliography is still controversial regarding this aspect. More properly conducted prospective studies have demonstrated higher frequencies of this complication, between 15 and 30%, related mainly to hyperglycemia in the pre-conception period (19).

According to the aforementioned article, the development of GDM occurs mainly at the end of pregnancy (15). Thus, the concentration of GDM diagnoses in the second and third semesters is explained, highlighting the incidence of 64% of GDM diagnoses in the third trimester. Regarding this, a descriptive retrospective study conducted in the same city, but in a public maternity ward, did not find an influence of the trimester of diagnosis on the occurrence of outcomes related to the type of treatment used and the classification of the newborn’s weight (20).

Regarding the glycemic profile of the studied patients, the influence of inadequate glycemic control on the incidence of newborns large for gestational age (LGA) is highlighted (21). In this sense, a research carried out from 2004 to 2006, in which 157 pregnant women presented as the most related factor to the presence of LGA newborns a two-hour glucose tolerance test of 75 g with a cut-off point of 170 mg/dL, in which values higher Increased the chance of LGA newborns by 3.7 times (22). In the present study, an average two-hour glucose tolerance test of 112.7 was found, considerably below the risk value, corroborating the importance of adequate prenatal care and correct glycemic control.

Similarly, the average value of glycated hemoglobin found was 5.26. Studies show an association of fetal malformation with third trimester fasting glycemia levels higher than 95 mg/dL and/or with glycated hemoglobin higher than 7.0 mg/dL (p of these associations of 0.04 and 0.05 respectively) (5).

Regarding the chosen treatment, 197 patients (38%) only underwent treatment for GDM with diet and physical activity. According to the 2019 Brazilian Consensus for GDM Treatment, it should be considered that, in Brazil, it is not always possible to count on a nutritionist for individual care of pregnant women with GDM, but the nutritional therapy recommendations presented should be implemented according to the financial and technical feasibility available in the locality (23). In this sense, an article that analyzed the most recent advances in GDM treatment concluded that the ideal standard of glycemic control begins with the union of multidisciplinary teams and technological improvement, but due to the high costs it is important to intensively seek alternative and accessible mechanisms to the entire population (25).

Regarding the newborn’s profile, the literature indicates diabetes mellitus, either gestational or pre-gestational, as the main risk factor for the gestation of a macrosomic fetus (25). In this sample we found an average weight of 3,123.4 g, within the normal parameters for the general population (26). In addition, the average gestational age of 37.4 weeks was also within the normality and the Apgar values at first and at fifth minute were also above 7, with no negative outcomes in the mean of the studied GDM population. However, this result does not exclude the incidence of individual outcomes, only the average of the data to find the profile of the GDM population in the service.

A perinatal outcome strongly related to GDM was the delivery route, since cesarean section corresponded to 81.4% of the patients studied. According to WHO data, Brazil has the second highest cesarean rate in the world with 55%, losing only to the Dominican Republic, where the rate is 56% (27). The literature indicates the high rate of cesarean as an important outcome of GDM (28,29), but the incidence found was even higher than the LINDA-Brasil cohort study (Lifestyle Intervention for Diabetes prevention After Pregnancy), which includes 3,325 pregnant women with GDM attended by the Unified Health System in Porto Alegre, Pelotas and Fortaleza and resulted in 64.7% of cesareans (30).

Regarding large for gestational-age (LGA) newborns, it was observed that 28.2% had this neonatal outcome. This illustrates a higher number than that found in the literature that currently indicates close to 20% (ref). On the other hand, in prematurity the result of this study (10.9%) was lower than that found in the literature (14.2%). However, these small variations in prematurity and LGA must be explained by population variants and by the fact that the present study was carried out in the private health service.

There are few recent studies that examine neonatal hyperbilirubinemia in gestations complicated by GDM, having encountered a rate of 25.6% in our work. Our results are discrepant in relation to recent studies such as the 2018 Italian STRONG study, in which the hyperbilirubinemia rate was 10.4%, and the 2019 Australian study, which reached 7.5%. Such results reinforce the need for more studies that solidify knowledge about the topic, there being, at the moment, no consensus regarding neonatal hyperbilirubinemia.

Both hypoglycemia and hyperbilirubinemia in gestations affected by diabetes are associated with neonatal comorbidities, including respiratory distress syndrome. (1) In an Australian study, there was an increased risk for respiratory distress in the presence of hyperbilirubinemia and hypoglycemia, including an increased risk of hyperbilirubinemia in the presence of hypoglycemia. (2) In the Italian STRONG study, 3.9% of respiratory distress was found with rates of 10.4% and 7.2% of hyperbilirubinemia and hypoglycemia, respectively. (3) In our study, there was presence of respiratory distress in 6.8% of the newborns, with presence of 25.6% of hyperbilirubinemia and 9.9% of neonatal hypoglycemia. Therefore, the high rate of respiratory distress could be associated with the higher rate of hyperbilirubinemia in the studied population.

Newborns of mothers with GDM are subject to various complications, sometimes requiring more intensive care. In our study, the rate of admission to the ICU was 7.4%. This value is similar to others found in the literature, being a study even in the same city (6%[1] and 5.[2]), but also disagrees with others, varying the need for ICU admission between 15%(3) and 23.5%(4). Such variation may stem from the difference in the quality of prenatal care, team preparation, among others, and the prevalence of local adverse outcomes.

Despite this, the fact that it is a study conducted exclusively in the private network makes the number of study patients limited. To better assess it would be of great value a study that was conducted in a multicenter way, and with coverage of the Unified Health System, in order to have a greater N, and having greater external validity.

The pregnant women diagnosed with GDM presented more cesarean deliveries compared to the normal population, and most of the diagnoses were made in the third trimester. Among the adverse outcomes related to newborns, there were more cases of hyperbilirubinemia, prematurity, fetal hypoglycemia and greater need for neonatal ICU.

## Data Availability

All data produced in the present work are contained in the manuscript.

## Funding

This study did not receive any funding

## Conflicts of interest

None declared.

## Ethics approval

The study was approved by the corresponding ethics committee by the number 77220417.7.0000.5366.

## Data availability

All data produced in the present work are contained in the manuscript.

## Notes

### Competing Interest Statement

The authors have declared no competing interest.

### Author Declarations

The project was approved under the number CAAE 77220417.7.0000.5366 by the Research Ethics Committee (CEP) of the University of the Region of Joinville - UNIVILLE, Joinville (SC), Brazil.

